# Cancer risk algorithms in primary care: can they improve risk estimates and referral decisions?

**DOI:** 10.1101/2021.06.25.21259541

**Authors:** Olga Kostopoulou, Kavleen Arora, Bence Pálfi

**Affiliations:** Imperial College London, Department of Surgery & Cancer

## Abstract

**Background:** Cancer risk calculators were introduced to clinical practice in the last decade, but they remain underused. We aimed to test their potential to improve risk assessment and 2-week-wait referral decisions.

**Methods:** 157 GPs were presented with 23 vignettes describing patients with possible colorectal cancer symptoms. GPs gave their intuitive risk estimate and inclination to refer. They then saw the risk score of an algorithm (QCancer was not named) and could update their responses. Half of the sample was given information about the algorithm’s derivation, validation, and accuracy. At the end, we measured their algorithm disposition.

**Results:** GPs changed their inclination to refer 26% of the time and switched decisions entirely 3% of the time. Post-algorithm decisions improved significantly vis-à-vis the 3% NICE threshold (*OR* 1.45 [1.27, 1.65], *p*<.001). The algorithm’s impact was greater where GPs had underestimated risk. GPs who received information about the algorithm had more positive disposition towards it. A learning effect was observed: GPs’ intuitive risk estimates became better calibrated over time, i.e., moved closer to QCancer.

**Conclusions:** Cancer risk calculators have the potential to improve 2-week-wait referral decisions. Their use as learning tools to improve intuitive risk estimates is promising and should be further investigated.

## BACKGROUND

Improving cancer outcomes in England is a national priority. In 2018, 55% of cancers were diagnosed at stages 1 and 2.^1^ NHS England aims to raise this to 75% by 2028 by improving the early diagnosis of cancer.^2^ GPs can use the two-week-wait (2WW) referral pathway if they suspect cancer; the patient is then seen by a specialist within a target of two weeks. It was recently demonstrated that the 2WW pathway is effective in improving cancer outcomes: higher use of the pathway was associated with lower mortality for common cancers and lower odds of late-stage diagnosis.^3^ However, large variability between practices in their use of the 2WW referral pathway^4^ means that it may not fulfil its full potential. The variability has partly been explained by the organisation of the local health services^5^ and partly by GP decision making.^6,7^ Discriminating patients who should be referred on the 2WW pathway from those who do not need to is difficult, especially where early cancers present with vague, non-specific symptoms that could easily be attributed to other conditions.^8^ Using cancer risk calculators could improve cancer referral decision making,^9^ by helping GPs identify at-risk patients and, thus, reduce diagnostic delay, while reassuring them about low-risk patients who do not require referral, and thus avoid overloading the healthcare system.

Cancer risk calculators are algorithms that calculate the probability that a patient with symptoms has a current, undiagnosed cancer. QCancer^10^ and RAT^11^ are two established cancer risk calculators which have been integrated with the electronic health record in some parts of primary care. Studies of the implementation of cancer risk calculators in clinical practice have had mixed results: a cohort study found an increase in the number of investigations ordered and cancers diagnosed after RATs were provided to practices;^12^ a cluster randomised trial found no impact of GP education resources, which included RATs, on time to diagnosis;^13^ and a qualitative study of GPs doing simulated consultations suggested distrust of QCancer when it conflicted with clinical judgement.^14^ Indeed, despite these tools being available in primary care for almost a decade and their potential to improve the earlier diagnosis of cancer, they remain an underused resource.^15^

The study reported here is the first in a planned series of studies aiming to investigate how cancer risk algorithms influence clinical risk assessments and referral decisions, and to identify ways to optimise their introduction and presentation. The study involved GPs responding to a series of clinical vignettes online. We investigated whether GPs change their referral decisions in response to an unnamed algorithm, if decisions improve, and what factors influence decision change, specifically:

- the provision of information about the algorithm, and
- the GP’s intuitive risk estimates in relation to the algorithm, i.e., underestimation vs. overestimation of risk.

Anecdotally, GPs are not routinely informed about how algorithms introduced in their electronic health record have been elicited and validated and how accurate they are. It is plausible to expect that such information would improve trust in the algorithm and lead to greater willingness to follow its advice and integrate its probabilities into one’s own risk assessment and referral decisions. We also expected that GPs would err on the side of caution, putting more importance on misses than false positive referrals, and thus be less willing to change a referral decision when the algorithm suggested that the patient’s risk was lower (vs. higher) than what they had initially thought. Furthermore, we investigated GPs’ disposition towards the algorithm and associations with GP demographics, prior attitudes towards cancer risk calculators, and decision confidence.

## METHOD

### Sample size

We powered the study to detect a small effect (*f*^2^=0.02) of the algorithm on referral decisions with alpha of 5% and power of 95% in a multiple linear regression. The G*Power software (v. 3.1.9.4) estimated that we would need at least 863 responses. To account for data clustering (each GP responding to 20 vignettes), we adjusted this number by the Design Effect (DE).^16^ This is calculated using the formula DE=1+(n–1)*ICC, where n is the cluster size (the 20 vignettes), and ICC is the intra-class correlation. We estimated the ICC from the pilot data to be 0.088. Thus, DE=2.68. We adjusted the number of participants required by multiplying the 863 required responses with the DE and dividing by the cluster size: (863*2.68)/20=116. Thus, we estimated that we needed to recruit a minimum of 116 GPs.

### Materials

#### The vignettes

We prepared 23 clinical vignettes, each having a different combination of risk factors, symptoms and signs related to colorectal cancer. To prepare the vignettes, we used QCancer (https://qcancer.org) and inserted risk factors and symptoms so that vignette risk ranged from 0.58% to 57.23% (mean 14.10%, SD 18.97, median 4.18). As well as creating some new vignettes, the majority were modified from those previously used in a study by the lead author.^7^

Each vignette described a hypothetical patient presenting in general practice. All vignettes started with a list of demographics and risk factors (name, sex, age, BMI, smoking and alcohol intake), followed by the presenting problem. One or more of the relevant risk factors and symptoms in QCancer (type 2 diabetes, family history of gastrointestinal cancer, weight loss, appetite loss, abdominal pain, rectal bleeding, change in bowel habit, constipation, and anaemia) were incorporated into the description.

Three of the vignettes were used for familiarisation purposes and no data were collected (Appendix 1). The remaining twenty were split into two sets of ten to be completed on two different days to minimise fatigue. We made sure that the range, median, mean and standard deviation of risk estimates were almost identical in the two sets. We also counterbalanced the sets across participants, so that each set was completed first and second an equal number of times. All materials were presented online on the Qualtrics platform (qualtrics.com).

### Procedure

An invitation email was sent to the 400 GPs in our database – a database of e-mails compiled by the lead author and consisting of previous study participants, all currently practising in England. The invitation email included a brief description of the study and outlined the benefits of participation: remuneration of £60 and personalised feedback. Those interested in participating could follow a link in the email, which took them to an expression-of-interest form, where they could enter their NHS email address and GP practice code.

The procedure is described in detail in Appendix 2. Briefly, GPs initially provided their demographics; rated their confidence in assessing patients with possible cancer; answered questions about their awareness of cancer risk algorithms, availability in clinical practice, and frequency of use; and rated their attitude towards cancer risk calculators.

Half of the participants were then randomly allocated to receive information about the study algorithm. We aimed to inform them of the algorithm’s derivation, validation, and accuracy (see Appendix 2, Box 1). The algorithm was never named as QCancer. We then measured their understanding of the information and trust of the algorithm. No information about the algorithm was provided to the other half of the sample.

Subsequently, all participants responded to three practice vignettes, which were designed to represent three levels of cancer risk – low (1%), medium (6%) and high (40%) – and thus help GPs calibrate their risk estimates. The ten vignettes of the first set were then presented in a random order. Each vignette was followed by three questions about the estimated risk of colorectal cancer, the confidence intervals around that risk estimate, and the likelihood of making a 2WW referral decision. Referral decisions were measured on a 5-point scale ranging from 1 ‘highly unlikely’ to 5 ‘highly likely’. The scale allowed respondents to choose ‘uncertainty’ (3, scale mid-point) and thus gave us the opportunity to measure how the algorithm impacted uncertainty.

After these three questions were answered, the vignette was presented again, this time with the algorithmic estimate. Participants were invited to answer the same three questions as before. They were reminded of their earlier responses to make sure that any discrepancy pre- and post-algorithm was due to the algorithm and not forgetting. We asked respondents who did not wish to change their earlier estimates to re-enter them, to make sure that lack of updating responses was not driven by laziness.

Twenty-four hours after completing the first set of 10 vignettes, participants were automatically sent a link to the second set. The procedure in the second study session was the same as in the first session. After completing the second set of vignettes, all participants answered seven questions about their opinions and experience of the algorithm in the study (Algorithm Disposition Questionnaire – Appendix 2). Finally, all participants were given the opportunity to comment on any aspect of the study, if they wished.

### Analyses

We aimed to measure the impact of our manipulation (algorithm information provided vs. not provided) and GPs’ over-vs. underestimation of risk on their referral decisions.

### Creation of variables

We created a dichotomous variable denoting the position of the GPs’ intuitive (i.e., the initial, pre-algorithm) risk estimates in relation to QCancer: overestimation (1) vs. underestimation (0). We excluded responses where intuitive estimates matched QCancer. To measure changes in risk estimates, we subtracted the final from the initial estimate, and signed the difference so that positive values indicated changes consistent with the algorithm (the final estimate was closer to the algorithm than the initial estimate) and negative values indicated changes inconsistent with the algorithm. Similarly, to measure changes in referral inclination, we subtracted the final from the initial response on the 1-5 scale and signed the difference so that positive values indicated changes consistent with the algorithm and negative values indicated changes inconsistent with the algorithm. For example, if the algorithm estimated a higher risk than the GP, who subsequently gave a higher value on the response scale, the raw difference of the two response values on the scale would be negative but the adjusted would be positive. We also created a simpler, dichotomous variable for referral inclination, indicating whether respondents moved from one point on the response scale to another: ‘change’ (1) vs. ‘no change’ (0).

To determine whether referral decisions improved post-algorithm, we created two dichotomous variables: decision appropriateness (appropriate vs. not appropriate) and time of decision (pre-vs. post-algorithm). We defined decision appropriateness using the NICE risk threshold of 3% (https://www.nice.org.uk/guidance/ng12/evidence/full-guideline-pdf-2676000277). Therefore, if GPs indicated that they were either likely or highly likely to refer a vignette with QCancer risk score ≥3%, the decision was classed as appropriate. Similarly, if they indicated that they were either unlikely or highly unlikely to refer a vignette with QCancer risk score <3%, the decision was classed as appropriate. Otherwise, it was classed as inappropriate.

### Regression models

All regression models were multilevel with random intercepts by GP and vignette, unless otherwise indicated. The regression tables are presented in Appendix 4, in the sequence that they appear in the text. First, we ran two empty regression models, one for risk estimate changes, the other for changes in referral inclination to measure the impact of the algorithm on these two behavioural measures. To measure whether changes in risk estimates were associated with changes in referral inclination, we regressed inclination changes on risk estimate changes. We repeated the analysis as a logistic regression, using the simpler, dichotomous variable for inclination changes (change vs. no change). We then regressed each referral inclination variable on the two predictors of interest (algorithm information and position of GPs’ intuitive estimates vis-à-vis QCancer). We also explored the contribution of other variables by subsequently adding them to these two regression models in a single step:

- GP demographics (gender, years in general practice),
- confidence when assessing patients with symptoms that might indicate cancer, and
- general attitude towards cancer risk calculators

Using logistic regression, we regressed decision appropriateness on time of decision. In one analysis, uncertain decisions were classed as inappropriate. We then repeated the analysis excluding uncertain decisions from the calculations. Finally, we explored whether any learning had taken place as a result of the QCancer score repeatedly presented after each vignette, by measuring whether GPs’ intuitive risk estimates improved over time, i.e., moved closer to QCancer. We defined improvement as a reduction in the difference between GPs’ intuitive risk estimates and QCancer. We used the absolute values of this difference to avoid situations where overestimation and underestimation cancelled each other out. We regressed this absolute difference on study session (1^st^ vs. 2^nd^); in a separate model, we regressed it on vignette order (1-20).

Finally, we explored predictors of algorithm disposition. Using simple linear regression, we regressed participants’ score on the Algorithm Disposition Questionnaire (ADQ score) on

- GP demographics (gender, years in general practice),
- confidence when assessing patients with symptoms that might indicate cancer,
- general attitude towards cancer risk calculators, and
- algorithm information (present vs. absent).

All analyses were carried out using Stata 17.0 and were confirmed in R (version 4.0.3).

## RESULTS

We recruited 150 fully qualified GPs and 7 GP trainees (ST3/4) from a total of 124 GP practices across England. The number of GPs working in same practice ranged from 1 to 5 (median 1). Most practices (101/124, 81%) were represented by one GP in the sample. The mean age of the sample was 44 years (SD 8.7) and 53.5% of the participants were female. Average experience was 14 years since qualification (SD 9, Median 12). Three GPs answered this question giving one or two-digit numbers rather than a year and were thus excluded from the calculation of experience. The mean number of clinical sessions per week was 5.9 (SD 1.9, Median 6). Most participants indicated that they were confident most of the time when assessing patients with symptoms that might indicate cancer (2380/3140, 76%), while a substantial minority were confident some of the time (620/3140, 20%).

### Awareness, frequency of use and attitudes towards cancer risk calculators

Most participants were aware of cancer risk calculators (108/157, 69%). However, only 47 GPs (29.9%) had cancer risk calculators available in the electronic health record at their practice, with QCancer being the most common (Table 1). The sample reported a generally positive attitude towards cancer risk calculators, with a mean of 5.99 on the response scale from very negative (1) to very positive (9) (SD 1.54). Table 1 presents the number of GPs who indicated that a specific type of cancer risk calculator was available in the electronic health record at their practice, and their attitude towards these calculators. Table 2 shows that where a calculator was known to be available, half of the respondents used it sometimes and a large minority (40%) never used it. Thus, out of a total of 157 participants, only 28 (18%) were actively using a cancer risk calculator either sometimes or always.

**Table 1.** Numbers (%) of GPs who indicated that they had access to cancer risk calculators at their practice and their attitude towards them, presented by type of cancer risk calculator available.

| Cancer risk calculator | GPs with access | Attitude towards risk calculators* (mean, SD) |
| --- | --- | --- |
| Qcancer | 26 (55%) | 5.08 (1.72) |
| C the Signs | 7 (15%) | 6.43 (1.27) |
| Qcancer & C the Signs | 9 (19%) | 6.11 (2.09) |
| Qcancer & RAT | 2 (4%) | 7 (2.83) |
| Other | 3 (6%) | 4 (2.65) |
| Total | 47 (100%) | 5.48 (1.88) |

\* *“In general, how do you feel about having cancer risk calculators in clinical practice?”* Response scale: “very negative” [1] to “very positive” [9].

**Table 2.**
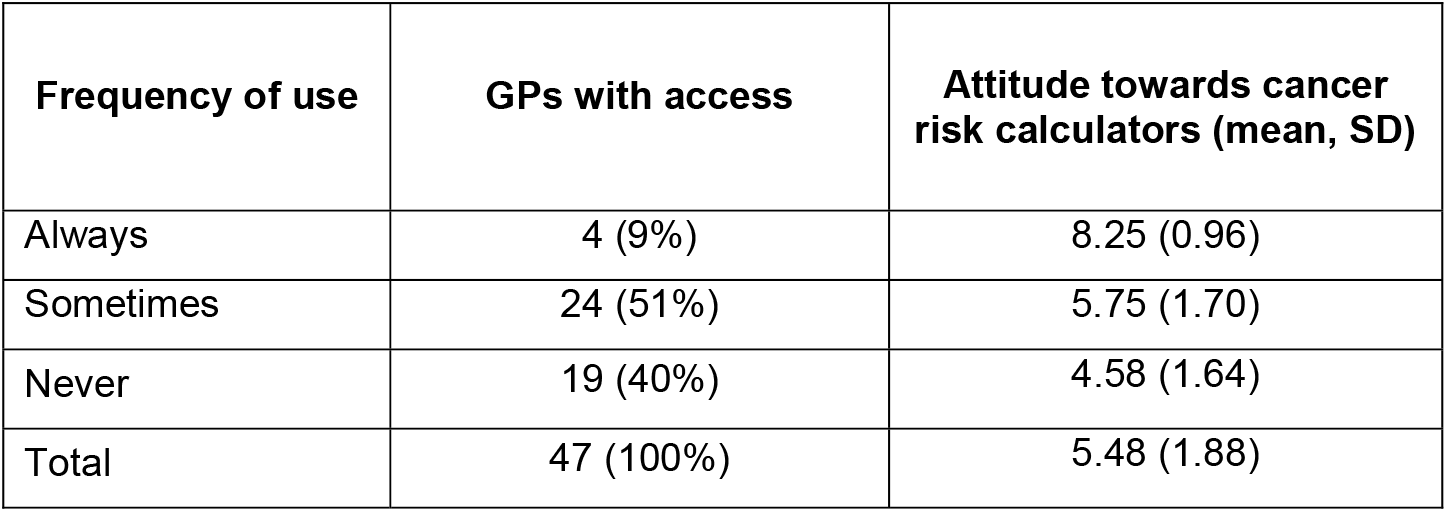
Frequency of cancer risk calculator use, where they were known to be available, and GPs’ attitude towards them.

| Frequency of use | GPs with access | Attitude towards cancer risk calculators (mean, SD) |
| --- | --- | --- |
| Always | 4 (9%) | 8.25 (0.96) |
| Sometimes | 24 (51%) | 5.75 (1.70) |
| Never | 19 (40%) | 4.58 (1.64) |
| Total | 47 (100%) | 5.48 (1.88) |

We provided half of our sample (80/157, 51%) with information about the algorithm at the start of the study (Appendix 2, Box 1). Most reported that the information made sense (78/80, 98%), that they would trust the algorithm’s estimates (“definitely yes” or “probably yes”: 72/80, 90%), and that they would like to have an algorithm like this in their clinical practice (“definitely yes” or “probably yes”: 72/80, 90%).

### Referral decisions pre- and post-algorithm

We collected a total of 3140 decisions about referral (157 participants responding to 20 vignettes). Table 3 summarises the moves on the decision response scale. Participants moved to a different point on the response scale after seeing the algorithm only in a quarter of responses (808/3140, 26%). In 12% of these responses, they switched their decisions entirely (95/808):

**Table 3.** Frequency of referral decisions pre- and post-algorithm. Decisions were measured on a 5-point scale ranging from 1 (highly unlikely) to 5 (highly likely), with a midpoint of 3 (uncertain).

| **Referral decisions** | **Pre-algorithm** | **Post-algorithm** |
| --- | --- | --- |
| Unlikely (1) or highly unlikely (2) | 545 (17.36%) | 637 (20.29%) |
| Uncertain (3) | 418 (30.67%) | 381 (32.42%) |
| Likely (4) or highly likely (5) | 2177 (69.33%) | 2122 (67.58%) |
| Total | 3140 (100%) | 3140 (100%) |

- from referral to no-referral (from 4 or 5 to 1 or 2 on the response scale) in 47 responses.
- from no-referral to referral (from 1 or 2 to 4 or 5 on the response scale) in 48 responses. In the remainder of responses (713/808), only the inclination to refer changed, either increasing (398/808, 49%) or decreasing (315/808, 39%).

### Changes in risk estimates and referral inclination and their association

Both risk estimates and referral inclination changed significantly after the algorithm was received; risk estimate changes: *b* = 10.23 [7.43, 13.03], *p* <.001; referral inclination changes: *b* = .25 [.20, .31] *p* <.001. The regression coefficients indicate that GPs changed their estimates in accordance with the algorithm by 10.23% on average and moved on the 1-5 decision response scale by a quarter of a unit on average. We found a weak but statistically significant association between changes in risk estimates and changes in referral inclination (*b* = .016 [.01, .02], *p* <.001).

We observed risk overestimation (intuitive estimate > QCancer score) in 70% of responses (2211/3140), underestimation in 23% of responses (714/3140), while intuitive estimates matched QCancer scores exactly in 7% of responses (215/3140). We categorised changes in referral inclination as either towards referral, i.e., any increase in value on the 1-5 response scale, or away from referral, i.e., any reduction in value on the 1-5 response scale. Table 4 shows that where GPs became more inclined to refer, they had underestimated risk on average and increased their risk estimates after seeing the algorithm; where they became less inclined to refer, they had overestimated risk on average and reduced their risk estimates after seeing the algorithm.

**Table 4.** Changes in the inclination to refer post-algorithm either towards or away from referral and associated means (SD) of GPs’ pre- and post-algorithm risk estimates and means (SD) of the QCancer risk score.

| Changes in inclination to refer |  | Risk estimate pre-algorithm | QCancer risk score | Risk estimate post-algorithm |
| --- | --- | --- | --- | --- |
| Towards referral | 327 (40.5%) | 12.4% (13.0) | 31.2% (22.6) | 27.5% (20.1) |
| Away from referral | 481 (59.5%) | 22.6% (20.0) | 3.5% (5.4) | 7.6% (9.5) |
| Total | 808 (100%) |  |  |  |

### Impact of algorithm information and position of intuitive risk estimates

Referral inclination changed in accordance with the algorithm more when risk was initially under-estimated vs. over-estimated (*b* = .31 [.24, .38], *p* <.001); we detected no effect of algorithm information. When we used the dichotomous decision variable in the regression, we found that the odds of change almost tripled when risk was initially under-estimated vs. over-estimated (*OR* = 2.84 [2.06, 3.90], *p* <.001); the test of algorithm information was not significant. Thus, it appears that GPs were being cautious and less willing to change their initial estimates and referral decisions when the algorithm suggested that they had overestimated cancer risk than when it suggested that they had underestimated it. Some GPs acknowledged this in their written comments. For example:

GP 12635: *“Low likelihood on the algorithm doesn’t really influence if I change my answers but a high likelihood does*.*”*

GP 27092: *“I think when the algorithm supported my decision, I found it helpful but when I would have thought to refer but it gave a low estimate, I often ignored it*… *If my own risk assessment was low but the algorithm’s was high, then I’d be more likely to err on the side of caution*.*”*

We repeated these regression analyses adding GP demographics, confidence when assessing possible cancers, and general attitude towards cancer risk calculators. We detected a significant negative relationship between changes in referral inclination and confidence when assessing patients with possible cancer symptoms (*b* = -.11 [-.20, -.02], *p* =.020). We also detected a significant positive relationship between the dichotomous variable (change vs. no change) and GPs’ general attitudes towards cancer risk calculators (*OR* = 1.18 [1.03, 1.35], *p* =.018).

### Algorithm impact on decision appropriateness

We categorised 63% of initial referral decisions and 68% of final referral decisions as appropriate; ‘uncertain’ responses were classed as inappropriate, in the first instance. GPs’ decisions post-algorithm were significantly more appropriate than pre-algorithm (*OR* = 1.45 [1.27, 1.65], *p* < .001). When we excluded from the count instances where either pre- or post-algorithm decisions were ‘uncertain’, the results were similar (*OR* = 1.26 [1.06, 1.50], *p* < .001, see Table 5 for frequencies). Thus, the odds of an appropriate referral decision were between 1.3 and 1.5 times higher after respondents received the algorithm’s risk estimates.

**Table 5.** Frequency of appropriate and inappropriate referral decisions before and after seeing the algorithm, with ‘uncertain’ responses first excluded and then included in the count as ‘inappropriate’.

|  | **'Uncertain' responses excluded**<br>**from count** |  | **'Uncertain' responses classed as**<br>**inappropriate** |  |
| --- | --- | --- | --- | --- |
|  | Pre-algorithm | Post-algorithm | Pre-algorithm | Post-algorithm |
| Appropriate | 1925 (75.2%) | 1984 (77.5%) | 1982 (63.1%) | 2147 (68.4%) |
| Inappropriate | 635 (24.8%) | 576 (22.5%) | 1158 (36.9%) | 993 (31.6%) |
| Total | 2560 (100%) | 2560 (100%) | 3140 (100%) | 3140 (100%) |

### Learning

We explored changes in intuitive risk estimates over time and observed the following:

1. The mean absolute difference between the GPs’ intuitive risk estimates and QCancer was significantly smaller in the second session: 17.3% (SD 16.83) in the first session vs. 15.70% (SD 15.91) in the second session. This difference between sessions was significant (b = -1.63% [-2.53, - 0.72] *p* < .001). Thus, during the second session, GPs closed the gap between their intuitive estimates and QCancer by 1.6 percentage points on average.
2. There was a significant negative relationship between vignette order and the absolute difference between the GPs’ intuitive risk estimates and QCancer (*b* = -.14 [-.22, -.06], *p* < .001), indicating that GPs’ estimates improved over time (moved closer to the algorithm). Figure 1 demonstrates this trend. The pattern of the results suggests that the improvement was not continuous but occurred mainly in the second session (vignette order 11 to 20). Furthermore, several GPs submitted comments that suggested awareness that learning had taken place, either in general or in relation to how specific symptoms contributed to a patient’s risk of colorectal cancer (Appendix 3).

**Figure 1.**
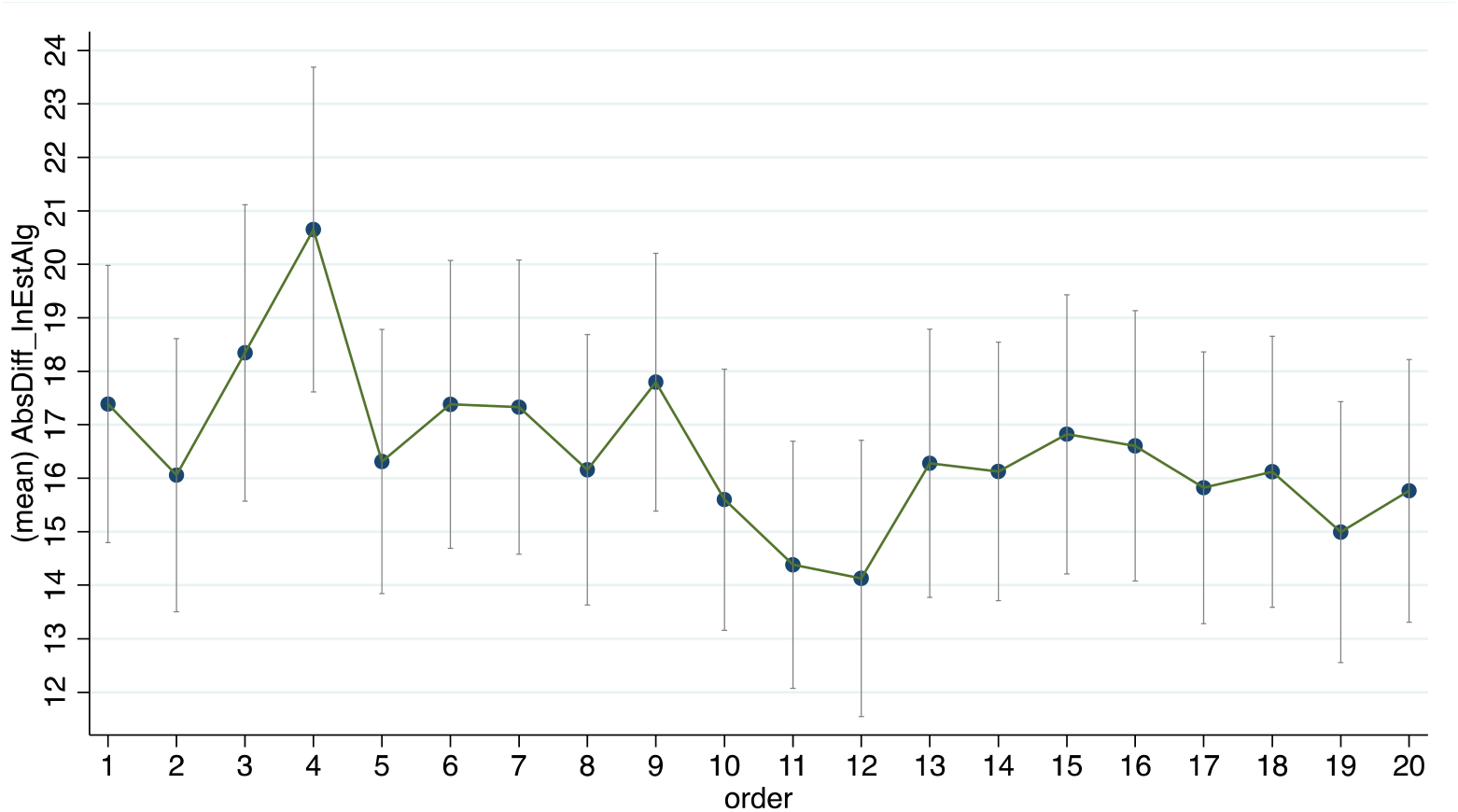
Line plot depicting the relationship between vignette order and the absolute difference between GPs’ intuitive risk estimates and QCancer scores. The dots represent the mean for each order and the error bars represent the 95% CIs.

### Algorithm Disposition Questionnaire (ADQ)

At the end of the study, GPs expressed a generally positive attitude towards the algorithm: the mean ADQ score was 5.11 – responses given on 7-point scales from strongly disagree (1) to strongly agree (7) (*SD* 1.13, *Median* 5.29). When we explored possible predictors of ADQ, we detected significant relationships with:

- gender (male = 0, female = 1): *b* = -.37 [-.73, -.02] *p* = .040,
- general attitude towards cancer risk calculators (*b* = .22 [.10, .33] *p* < .001),
- confidence when assessing patients with possible cancer symptoms (*b* = -.54 [-.91, -.18] *p* = .004), and
- algorithm information: *b* = .36 [.01, .70] *p* = .043.

## DISCUSSION

Our study findings provide insights into the potential impact and benefits of using cancer risk calculators in clinical practice for the earlier detection of cancer. In general, changes in inclination to refer were consistent with changes in risk estimates. Importantly, we measured an improvement in the appropriateness of referral decisions post-algorithm. Although this improvement was small, it was statistically significant. There are several reasons for the small effect of the algorithm on referral decisions. First, decisions are notoriously difficult to change, despite changes in associated risk estimates.^17^ Decisions may be based on other factors, in addition to or irrespective of risk estimates.^18,19^ For example, we found some evidence that both general attitudes towards cancer risk calculators and confidence in assessing patients with possible cancer symptoms were related to shifts in inclination to refer post-algorithm. Second, GPs abided by the precautionary principle: although around half of the vignettes had risk ≤3%, not necessitating referral, referrals post-algorithm remained high at 68%. Referral inclination changed in accordance with the algorithm more if GPs had underestimated than overestimated risk. In other words, GPs erred on the side of caution and were disinclined to change an initial referral decision, even if the algorithm indicated that referral was not required. Finally, decision appropriateness started from a relatively high baseline. GPs were already making good referral decisions pre-algorithm. Still, a 5% absolute improvement post-algorithm could translate into cancers diagnosed earlier, as well as unnecessary referrals being avoided.

GPs overestimated cancer risk most of the time. This may be because they do not regularly provide explicit risk estimates in the form of numerical probabilities. Risk estimates often remain implicit and GPs do not receive any systematic feedback that would enable them to calibrate their estimates better. Ours is not the only study that identified poor calibration of risk estimates. A survey of Canadian GPs found that they overestimated the absolute 8-year cardiovascular disease risk for two hypothetical patients.^20^ A recent study found that US clinicians at outpatient clinics overestimated the probability of disease for four scenarios common in primary care and that overestimation persisted after receiving both positive and negative test results.^21^ Focusing on single disorders in the absence of a differential in the above studies is also likely to have contributed to overestimation. Consideration of alternative diagnostic possibilities is one strategy to drive down overestimation.^22^

An unexpected but encouraging finding from our study was the learning effect, which became apparent during the second session: GPs became better calibrated, i.e., their intuitive risk estimates moved closer to the algorithm. This suggests that GPs were noticing and learning the probabilistic relationships between single or multiple symptoms and algorithmic risk. GPs were aware of this learning, as is evident in their comments. The finding that improvement occurred in the second session is consistent with the literature on learning consolidation, i.e., the stabilisation of memory traces after their initial acquisition.^23^ Consolidation of learning happens during off-line periods, when participants are not engaged with the task at hand. A fruitful avenue for future research would therefore be to explore the learning that occurs during repeated trials with algorithm feedback, and the factors affecting consolidation, such as the length of the time interval between training sessions, and the frequency and spacing of booster training sessions. Thus, risk calculators may have a role as training tools, enabling GPs to internalise the weighting of risk factors such as age, smoking, alcohol intake, family history, and specific symptoms on the risk of undiagnosed cancer in patients. GPs who participated in our study frequently commented on the opportunity for reflection that the study provided. Opportunities for reflection, learning and improvement could engender positives attitudes towards risk calculators.

We found moderately high awareness and a generally positive attitude towards cancer risk calculators amongst participants at the start of the study. Nevertheless, the reported availability in clinical practice and the self-reported use of these calculators was low. Our study confirms that cancer risk calculators remain an underused resource. A cross-sectional postal survey of UK GPs in 2017 also found low awareness, availability and use of cancer risk calculators.^15^ However, the positive comments about the algorithm made by our participants at the end of the study suggest that barriers to the adoption of cancer risk calculators are most likely practical, such as a lack of supportive activities during their introduction^24^ and lack of seamless integration into the clinical workflow.^14^ External support from cancer networks in the form of webcasts, email updates and newsletters were found to improve the acceptance and use of RATs.^24^ Therefore, a combination of external support and training sessions may help to increase the uptake of cancer risk calculators in the future.

Our study also aimed to investigate whether informing GPs of the algorithm’s provenance, validation and accuracy increases their willingness to change their decisions in accordance with the algorithm. We did not find evidence for this. We did however find that GPs who received information about the algorithm expressed more positive attitudes towards it at the end of the study. We therefore recommend that risk algorithms are always introduced with care in clinical practice, ensuring that users have all the necessary information about them.

To study the impact of the algorithm on clinical judgements and decisions, we created a controlled environment quite different from real-life clinical consultations. Our participants saw a series of patient descriptions in sequence, all containing features and risk factors associated with colorectal cancer, and they were specifically asked to consider the possibility of colorectal cancer. In practice, these patients are unlikely to present one after the other or on the same day. They may present with a multitude of vague symptoms that make their diagnosis and management more uncertain. GPs may be considering other diagnoses, in addition to or perhaps at the exclusion of colorectal cancer, and they may elicit different information. They will elicit information in sequence and not in a neat, brief summary as presented in the study. They may order investigations – not provided in the study, and they may ask the patient to come back for a follow-up consultation. They may need to deal with external pressures, such as patient expectations, or practice policies to increase or reduce cancer referrals. They will not be asked to provide explicit risk estimates and, if they wish to make a decision about referral, they will not make it on a 1-5 response scale. One can think of a multitude of differences between responding to concise written descriptions of patients on computer and dealing with real people at a busy practice. We had to control the amount, type and format of information provided to participants to ensure standardisation and remove confounders. Instead of a dichotomous yes/no referral decision, and mindful that our participants would sometimes need more information or a further consultation before making a decision, we opted for a response scale that measured inclination rather than a final decision and offered them the option to remain ‘uncertain’. We do not claim that our GPs would respond in the same way to these patients if they saw them at their practice – even though, there is evidence that responding to written clinical vignettes provides a good approximation of real-life behaviour.^25^ Our aim was to determine to what extent GPs are willing and able to integrate estimates from a cancer risk calculator into their own intuitive estimates and if such calculators have the potential to improve referral decisions. Our findings provide a positive answer to both questions.

Our study demonstrates the potential for more appropriate 2WW referrals to be made if cancer risk calculators are used routinely. However, 2WW referral recommendations are currently underpinned by NICE guidance with no requirement for explicit risk scores to be determined prior to referral. This lack of integration of cancer risk calculators in the NICE referral criteria may cause GPs to question the incentives of using a risk calculator, which in practical terms does not supersede NICE guidance. Indeed, until clinical scoring systems are adopted into guidelines, their routine use in clinical practice is limited.^26^ The role of cancer risk calculators in clinical practice needs to be clarified for GPs, so that their advantages can be maximised. Risk scores could be included in referral criteria and referral forms, providing a backup for GPs to refer in non-specific cases, where NICE guidance is not met but the score fulfils the recommended risk threshold for referral. There may be value in including risk scores in referrals to vague symptom clinics or rapid access diagnostic clinics for non-specific presentations. For example, one such pathway, the Suspected CANcer (SCAN) in Oxfordshire which opened in 2017, asks GPs to enter their intuitive cancer risk estimate but has no place for a calculated risk score.^27^ Another potential purpose for cancer risk calculators is for counselling patients, providing reassurance when the risk is low and encouraging action where patients are reluctant to be investigated.^14^ Making patients aware of these calculators may further encourage their use during the consultation.

In summary, there is value in the use of cancer risk calculators in clinical practice, but they are currently underused. Their potential role as resources for training should be further explored and they could become part of training materials for GP trainees and new GP starters. The desired result would be a better uptake of these tools, as well as a greater understanding of the weighting of risk factors and symptoms when assessing patients, with the ultimate aim to improve the early diagnosis of cancer.

## Supporting information

Three of the vignettes were used for familiarisation purposes and no data were collected (Appendix 1)

## Data Availability

The dataset will be deposited on Dryad and will be publicly available upon acceptance for journal publication.

## Author contributions

OK conceived the study, obtained the funding, and acted as project leader. All authors contributed to the original idea for the study. OK and KA drafted the paper. KA adapted and created clinical vignettes with input from OK and BP. BP programmed the vignette survey on the Qualtrics platform and collected the data. OK and BP conducted the statistical analyses. All authors discussed the content and contributed to editing the manuscript.

## Ethics approval and consent to participate

Study approval was provided by the Health Research Authority (HRA) and Health & Care Research Wales (HCRW), REC reference 20/HRA/2418.

Participants provided consent online after reading a Participant Information Sheet. If they decided to withdraw from the study at any point, they could close the browser, in which case their data would be deleted.

The study was performed in accordance with the Declaration of Helsinki.

## Data availability

The dataset will be deposited on Dryad and will be publicly available upon acceptance.

