## Supplementary material for "Cancer risk algorithms in primary care: can they improve risk estimates and referral decisions?": Three of the vignettes were used for familiarisation purposes and no data were collected (Appendix 1)

Affiliation for all authors: Imperial College London, Department of Surgery & Cancer

**ABSTRACT**

**Background:** Cancer risk calculators were introduced to clinical practice in the last decade, but they remain underused. We aimed to test their potential to improve risk assessment and 2-week-wait referral decisions.

**Methods:** 157 GPs were presented with 23 vignettes describing patients with possible colorectal cancer symptoms. GPs gave their intuitive risk estimate and inclination to refer. They then saw the risk score of an algorithm (QCancer was not named) and could update their responses. Half of the sample was given information about the algorithm’s derivation, validation, and accuracy. At the end, we measured their algorithm disposition.

**Results:** GPs changed their inclination to refer 26% of the time and switched decisions entirely 3% of the time. Post-algorithm decisions improved significantly vis-à-vis the 3% NICE threshold (*OR* 1.45 [1.27, 1.65], *p*<.001). The algorithm’s impact was greater where GPs had underestimated risk. GPs who received information about the algorithm had more positive disposition towards it. A learning effect was observed: GPs’ intuitive risk estimates became better calibrated over time, i.e., moved closer to QCancer.

**Conclusions:** Cancer risk calculators have the potential to improve 2-week-wait referral decisions. Their use as learning tools to improve intuitive risk estimates is promising and should be further investigated.

**Supplementary material (Appendices)**

**Appendix 1: Example vignettes**

**Patient name: Bryony Barnes (female)**

**Age: 56**

**BMI: 30**

**Smoking: Currently smokes 15 cigarettes/day**

**Alcohol intake: 21 units/week**

Bryony Barnes comes to see you complaining of being more constipated in the last month. Over the last 2 months, she has also noted that she has lost about 4kg in weight and doesn’t understand why. She has not been dieting and her lifestyle has not changed.

**QCancer risk score: 1.04%**

**Patient name: Henry Lipp (male)**

**Age: 75**

**BMI: 24.9**

**Smoking: Never smoked**

**Alcohol intake: 4 units/week**

Mr Henry Lipp has come to see you concerned because he noticed some blood in his stools over the last four weeks. He has no other symptoms.

**QCancer risk score: 6.33%**

**Patient name: Dawn Jenkins (female)**

**Age: 70**

**BMI: 26.2**

**Smoking: Ex-smoker**

**Alcohol intake: 14 units/week**

Dawn Jenkins is your next patient. She has a background of Type 2 Diabetes and no other medical problems. She has come to see you because in the last few weeks she has become increasingly aware of some right sided abdominal pain. When you ask her about her bowels, she says she has seen some blood on and off when she looks in the toilet and this has been the case for the last two weeks. You ask her to do some blood tests which reveal a microcytic anaemia (Hb 9.8) and a low ferritin.

**QCancer risk score: 39.60%**

**Appendix 2. Study procedure**

After participants accessed the study site, they read an information sheet and provided consent online. They then completed questions about demographics (age, gender, GP or GP trainee, year of GP qualification, number of clinical sessions per week) and rated how often they felt confident when assessing patients with symptoms that might indicate cancer (always / most of the time / sometimes / seldom). They then answered questions concerning their awareness of cancer risk algorithms, availability in clinical practice, and frequency of use:

- *Are you aware of any cancer risk algorithms that are being used in clinical practice to calculate a patient’s current risk of cancer (aka 'cancer risk calculators')?* (Yes / No)

If the answer to the above question was ‘yes’,

- *Are they available in the electronic health record that you use in your practice?* (Yes / No)

If the answer to the above question was ‘yes’,

- *Which of the following (if any) cancer risk algorithms do you have in your practice?* (RAT / QCancer / C the Signs / Other) [more than one option could be chosen]
- *How often do you use them?* (always / sometimes / never)

Finally, all participants were asked to rate their attitude towards cancer risk calculators:

- *“In general, how do you feel about having cancer risk calculators in clinical practice?”* Very negative (1) – Very positive (9)

Half of the participants were randomly allocated to receive the following information about the study algorithm:

The algorithm aims to be used as a decision aid, to support 2WW cancer referral decisions. It is not intended to determine those decisions.

The algorithm was derived from a large cohort study of 2.5 million patients in the UK. They used data in the primary care record of cancer patients to estimate associations between risk factors, symptoms/signs and a subsequent cancer diagnosis.

The algorithm estimates the probability that a patient has colorectal cancer, given his/her risk factors and presenting symptoms/signs; in other words, how many people out of 100 with the same risk factors and presenting symptoms/signs are likely to have colorectal cancer.

A study that validated the algorithm on another large cohort of patients, a proportion of whom had colorectal cancer, found that the algorithm performed very well: it discriminated correctly between cancer and non-cancer patients approximately 90% of the time (i.e., produced higher risk estimates for cancer than non-cancer patients).

**Box 1. Algorithm description**

Participants in the algorithm information group then responded to three questions gauging understanding and trust:

1. Does the description above make sense to you? (Yes / No)
2. Would you trust the estimates of this algorithm? (Definitely yes / Probably yes / Probably not / Definitely not)
3. Would you like to have an algorithm like this in your clinical practice? (Definitely yes / Probably yes / Probably not / Definitely not)

All participants were then presented with the three practice vignettes in a random order. No data were collected at this stage and participants were informed of this. The aim of the practice vignettes was to familiarise participants with the task and help them calibrate their risk estimates, since GPs do not provide explicit cancer risk estimates on a routine basis. For this purpose, the practice vignettes represented three levels of risk of undiagnosed colorectal cancer: low (1%), medium (6%) and high (40%).

The ten vignettes of the first set then followed in a random order. The procedure was exactly the same for all the vignettes, including the practice ones. Specifically, each vignette was followed by three questions:

1. “Out of 100 patients with the same risk factors and symptoms as this patient, how many, in your clinical judgement, are likely to have colorectal cancer? Please type in a whole number between 0 and 100.”

Responses could be typed in a box below the question.

2. “What is the narrowest range which you are almost certain contains your estimate above? Enter the lower and upper limits in the boxes below. Make sure that your estimate falls within this range.

*I am almost certain that out of* ***100*** *patients like this one, between <the lower limit> and <the upper limit> are likely to have colorectal cancer as yet undiagnosed.*

Responses could be typed in two boxes, one for the lower limit, the other for the upper limit.

3. “How likely is it that you would refer this patient on the 2WW pathway for suspected cancer **at this consultation**?”

Responses were given on a 5-category response scale by selecting between ‘Highly Unlikely’, ‘Unlikely’, Uncertain’, ‘Likely’, and ‘Highly likely’

NB. Words in bold or italics appeared on the screen as they appear below.

After these three questions were answered, the vignette was presented again, this time with the algorithmic estimate: *“The algorithm estimates that <number> out of 100 patients presenting like this is likely to have colorectal cancer. Your estimate was <number> out of 100 (lower limit <number>, upper limit <number>). If you wish to revise your initial estimates, please do so below. If you wish to stick with your initial estimates, please re-enter them below.”* Participants were then invited to answer the same three questions as before.

Following completion of the first 10 vignettes, participants had the opportunity to give feedback on any aspect of the study in free text. Twenty-four hours after completing the first set of 10 vignettes, participants were automatically sent a link to the second set. The procedure in the second study session was the same as in the first session. Participants who had received information about the algorithm in the first session, were presented with it again at the start of the second session. After completing the second set of vignettes, all participants completed the Algorithm Disposition Questionnaire (ADQ).

- I found the algorithm’s risk estimates helpful.
- I think that the algorithm’s estimates were accurate.
- I felt irritated when receiving the algorithm’s estimates. (reverse-scored)
- I was happy to receive the algorithm’s estimates.
- I was frustrated when receiving the algorithm’s estimates. (reverse-scored)
- I felt more confident in my referral decisions, having received the algorithm’s estimates.
- I feel appreciative having access to the algorithm’s estimates.

Answers to each question were given on 7-point scales:

1 (strongly disagree), 2 (disagree), 3 (slightly disagree), 4 (neither disagree nor agree), 5 (slightly agree), 6 (agree), 7 (strongly agree)

Finally, all participants were given the opportunity to comment on any aspect of the study, if they wished.

**Appendix 3: Comments made by GPs at the end of the study in relation to reflecting and learning from the algorithm**

GP 20192: *Really enjoyable and thought provoking, made me question and refine my decision making…I felt an element of learning not just problem solving.*

GP 26347: *I would have liked to have a practice at this first before submitting to get used to the idea of algorithms and the statistics, as this improved during the course of this exercise.*

GP 43463: *Very realistic cases. Definite educational value when going through a list of cases like this.*

GP 74216: *Useful exercise, supporting reflection on one’s own assumptions.*

GP 78939: *Good cases, slightly difficult to put a number to symptoms but got easier as I went along.*

GP 95191: *I think this would make an interesting teaching resource.*

GP 78939: *Some of the algorithm results for the cases surprised me in that I wouldn’t have had the risk that high, so good learning points to consider…Good cases, slightly difficult to put a number to symptoms but got easier as I went along.*

GP 17080: *Very interesting how the algorithm generally estimated much lower than my personal judgement. My personal estimation of likelihood of cancer also appeared relatively 'blunt' (generally just 'high') compared to the algorithm.*

Specific comments about learning how symptoms contributed to cancer risk:

GP 97506: *That was a really useful way to learn about what factors the algorithm weights. Thank you for the opportunity to take part. Feels like the results of this study will be really clinically relevant for day-to-day practice!*

GP 25029: *I think I underestimate the effect of obesity and alcohol when I assess patients! Interesting to see on these patients my internalised score vary most widely.*

GP 29594: *Interesting experience thanks - will make me note abdominal pain as a symptom more.*

GP 61790: *In some cases, the alcohol intake, smoking status or BMI appear to have a significant impact on the algorithm estimate of probability of colorectal cancer. This is helpful.*

GP 95191: *Seemed age/smoking influenced things quite strongly.*

**Appendix 4: Regression tables**

**Changes in risk estimates and referral inclination and their association**

| **DV: Risk estimate changes** | **b** | **SE** | **t** | **P** | **95% CI** |
| --- | --- | --- | --- | --- | --- |
| Constant | 10.23 | 1.43 | 7.16 | <0.001 | 7.43, 13.03 |

Table S1. Empty multilevel linear regression model measuring the extent of change in risk estimates post-algorithm

| **DV: Referral inclination changes** | **b** | **SE** | **t** | **P** | **95% CI** |
| --- | --- | --- | --- | --- | --- |
| Constant | 0.25 | 0.03 | 8.59 | <0.001 | 0.20, 0.31 |

Table S2. Empty multilevel linear regression model measuring the extent of change in the inclination to refer post-algorithm

| **DV: Referral inclination changes** | **b** | **SE** | **t** | **P** | **95% CI** |
| --- | --- | --- | --- | --- | --- |
| Risk estimate changes | 0.016 | 0.00 | 18.08 | <0.001 | 0.01, 0.02 |
| Constant | 0.09 | 0.02 | 2.89 | 0.006 | 0.03, 0.15 |

Table S3a. Multilevel linear regression measuring the association between change in risk estimates and change in the inclination to refer post-algorithm.

| **DV: Referral inclination change (binary)** | **OR** | **z** | **P** | **95% CI** |
| --- | --- | --- | --- | --- |
| Risk estimate changes | 1.05 | 10.86 | <0.001 | 1.04, 1.05 |
| Constant | 0.16 | 11.25 | <0.001 | 0.12, 0.22 |

Table S3b. Multilevel logistic regression measuring the association between change in risk estimates and change in referral inclination as a binary variable.

**Impact of algorithm information and position of intuitive risk estimates**

| **DV: Referral inclination changes** | **b** | **SE** | **t** | **P** | **95% CI** |
| --- | --- | --- | --- | --- | --- |
| Position of intuitive risk estimates |  | | | | |
| Underestimation | 0.31 | 0.04 | 8.87 | <0.001 | 0.24, 0.38 |
| Algorithm information |  | | | | |
| Provided | 0.05 | 0.04 | 1.13 | 0.261 | -0.03, 0.13 |
| Constant | 0.22 | 0.04 | 5.56 | 0.000 | 0.14, 0.39 |

Table S4a. Multilevel linear regression measuring the impact of algorithm information (provided vs. not provided) on changes in referral inclination, and the association with risk under- vs. over-estimation.

| **DV: Referral inclination change (binary)** | **OR** | **z** | **P** | **95% CI** |
| --- | --- | --- | --- | --- |
| Position of intuitive risk estimates |  | | | |
| Underestimation | 2.84 | 6.39 | <0.001 | 2.06, 3.90 |
| Algorithm information |  | | | |
| Provided | 1.09 | 0.41 | 0.680 | 0.73, 1.63 |
| Constant | 0.21 | -8.33 | <0.001 | 0.15, 0.30 |

Table S4b. Multilevel logistic regression measuring the impact of algorithm information (provided vs. not provided) on changes in referral inclination as a binary variable, and the association with risk under- vs. over-estimation.

| **DV: Referral inclination changes** | **b** | **SE** | **t** | **P** | **95% CI** |
| --- | --- | --- | --- | --- | --- |
| Position of intuitive risk estimates |  | | | | |
| Underestimation | 0.31 | 0.04 | 8.46 | <0.001 | 0.24, 0.39 |
| Algorithm information |  |  |  |  |  |
| Provided | 0.04 | 0.04 | 0.89 | 0.377 | -0.05, 0.12 |
| GP gender |  | | | | |
| Female | -0.05 | 0.05 | -1.21 | 0.227 | -0.14, 0.03 |
| GP experience | 0.002 | 0.00 | 0.92 | 0.358 | -0.002, 0.01 |
| Decision confidence | -0.11 | 0.05 | 2.35 | 0.020 | -0.20, -0.02 |
| Attitude towards risk calculators | 0.02 | 0.01 | 1.62 | 0.108 | -0.005, 0.052 |
| Constant | 4.72 | 4.74 | 1.00 | 0.321 | -4.57, 14.02 |

Table S5. Extension of the multilevel linear regression model reported in Table S4a above, by adding GP demographics (gender and experience), general confidence in assessing possible cancers and general attitude towards cancer risk calculators.

| **DV: Referral inclination change (binary)** | **OR** | **z** | **P** | **95% CI** |
| --- | --- | --- | --- | --- |
| Position of intuitive risk estimates |  | | | |
| Underestimation | 2.76 | 6.07 | <0.001 | 1.83, 4.16 |
| Algorithm information |  | | | |
| Provided | 1.04 | 0.17 | 0.869 | 0.69, 1.56 |
| GP gender |  | | | |
| Female | 0.85 | -0.77 | 0.442 | 0.55, 1.30 |
| GP experience | 1.01 | 1.12 | 0.261 | 0.99, 1.03 |
| Decision confidence | 0.67 | -1.80 | 0.072 | 0.44, 1.04 |
| Attitude towards risk calculators | 1.18 | 2.36 | 0.018 | 1.03, 1.35 |
| Constant | 4.31*10^8^ | 1.04 | 0.297 | 2.66*10^-8^, 6.97*10^24^ |

Table S5b. Extension of the multilevel logistic regression model reported in Table S4b above, by adding GP demographics (gender and experience), general confidence in assessing possible cancers and general attitude towards cancer risk calculators.

**Algorithm impact on decision appropriateness**

| **DV: Decision appropriateness** | **OR** | **SE** | **z** | **P** | **95% CI** |
| --- | --- | --- | --- | --- | --- |
| Time of decision |  | | | | |
| Post-algorithm | 1.45 |  | 5.55 | <0.001 | 1.27, 1.65 |
| Constant | 3.38 |  | 3.02 | 0.003 | 1.53, 7.45 |

Table S6a. Multilevel logistic regression of decision appropriateness on time of decision (pre- vs. post-algorithm). The ‘uncertain’ responses on the 1-5 scale have been excluded from the calculation.

| **DV: Decision appropriateness** | **OR** | **SE** | **z** | **P** | **95% CI** |
| --- | --- | --- | --- | --- | --- |
| Time of decision |  | | | | |
| Post-algorithm | 1.26 |  | 2.67 | 0.008 | 1.06, 1.50 |
| Constant | 6.77 |  | 4.10 | <0.001 | 2.71, 16.90 |

Table S6b. Multilevel logistic regression of decision appropriateness on time of decision (pre- vs. post-algorithm). The ‘uncertain’ responses on the 1-5 scale have been included in the model and categorised as ‘inappropriate’.

**Learning**

| **DV: \|Intuitive estimate – algorithm\|** | **b** | **SE** | **z** | **P** | **95% CI** |
| --- | --- | --- | --- | --- | --- |
| Study session |  | | | | |
| Second | -1.63 |  | -3.51 | <0.001 | -2.53, -0.72, |
| Constant | 17.32 |  | 8.49 | <0.001 | 13.32, 21.31 |

Table S7. Multilevel linear regression of the absolute difference between risk estimates pre-algorithm and QCancer risk scores by study session (first vs. second).

| **DV: \|Intuitive estimate – algorithm\|** | **b** | **SE** | **t** | **P** | **95% CI** |
| --- | --- | --- | --- | --- | --- |
| Vignette order | -0.14 | 0.04 | 8.69 | <0.001 | -0.22, -0.06 |
| Constant | 18.00 | 2.07 | 8.69 | <0.001 | 13.94, 22.06 |

Table S8. Multilevel linear regression of the absolute difference between risk estimates pre-algorithm and QCancer risk scores on vignette order (1-20).

**Algorithm Disposition Questionnaire (ADQ)**

| **DV: ADQ score** | **b** | **SE** | **t** | **P** | **95% CI** |
| --- | --- | --- | --- | --- | --- |
| GP gender |  | | | | |
| Female | -0.37 | 0.18 | -2.07 | 0.040 | -0.73, -0.02 |
| GP experience | -0.01 | 0.01 | -0.86 | 0.392 | -0.03, 0.01 |
| Decision confidence | -0.54 | 0.19 | -2.93 | 0.004 | -0.91, -0.18 |
| Attitude towards risk calculators | 0.22 | 0.06 | 3.69 | <0.001 | 0.10, 0.33 |
| Algorithm information |  | | | | |
| Provided | 0.36 | 0.17 | 2.04 | 0.043 | 0.01, 0.70 |
| Constant | 5.49 | 0.67 | 8.19 | <0.001 | 4.17, 6.82 |

Table S9. Simple linear regression of the ADQ score on GP demographics (gender and experience), general confidence in assessing possible cancers, general attitude towards cancer risk calculators, and algorithm information.
